# Effects of morning and evening narrowband blue light and myopic defocus on axial length in humans

**DOI:** 10.64898/2026.03.03.26347502

**Authors:** Swapnil Thakur, Harnish Khudkhudia, Padmaja Sankaridurg, Pavan K. Verkicharla

## Abstract

**Purpose:** To investigate the effects of morning and evening narrowband blue light exposure on axial length, and to examine the short-term effect of morning blue light combined with myopic defocus on axial length.

**Methods:** For objective 1, 18 individuals underwent 60 minutes of narrowband blue light exposure (460nm) in the morning (9:00–11:00AM) and evening (5:00–7:00PM) of the same day. The axial length values were normalized to the average of the morning and evening axial length values. For objective 2, 27 young adults were exposed to 60 minutes of narrowband blue light and broadband white light while wearing a +3.00 D lens over the right eye. Axial length was measured using Lenstar LS900.

**Results:** A significant reduction in axial length was observed after exposure to morning blue light compared to evening blue light (−10.0±3.96µm vs.−0.67±3.30µm; p=0.02), whereas no such effect was observed with broadband white light exposure (0.0±3.53 µm vs. -2.50±4.23µm, p=0.70). While the broadband white light exposure did not alter the normal diurnal variation in axial length (+2.35±1.82µm vs.–6.25±2.21µm, p=0.04), blue light diminished such a pattern (-4.12±1.72µm vs. - 2.00±2.00µm, p=0.48). The myopic defocus did not influence axial length under either narrowband blue or broadband white light conditions.

**Conclusion:** The short-term narrowband blue light exposure led to a significant decrease in axial length in the morning than evening exposure, with a likely influence on the diurnal rhythm of axial length. Morning blue light exposure with lens-induced myopic defocus did not provide additional short-term modulation of axial length.

## Introduction

Exposure to narrowband short-wavelength light has received increasing attention in the context of circadian regulation. Recent evidence from several studies assessed through a systematic review revealed that daytime exposure to narrowband blue light plays a key role in suppressing melatonin and enhancing alertness, whereas chronic exposure to even low-intensity blue light before bedtime may adversely shift the circadian clock.^1^ Furthermore, animal studies demonstrate that exposure to specific narrowband wavelengths can modulate ocular growth and refractive development, although these effects vary across species. For example, short-wavelength light (violet, blue) inhibits axial elongation in mice, chickens, fish, and guinea pigs,^2–11^ while in tree shrews and rhesus monkeys, red light inhibited axial elongation. In contrast, zebrafish show reduced ocular growth under both cyan and red light.^12^ Nickla et al.^10^ examined the interaction of narrowband blue light exposure with the intrinsic ocular diurnal rhythm in chicks and found that morning exposure to blue light results in shorter ocular axial dimensions compared to evening exposure, and that narrowband blue light disrupted the ocular rhythm. More recently, Liu et al.^13^ demonstrated that human participants using blue-pass filters for a shorter duration exhibited relatively shorter axial lengths during the daytime, suggesting an alteration of the diurnal rhythm, which is usually characterized by a physiologically longer axial length during daytime and a shortening phase later in the day.^14–16^ Collectively, these findings suggest that narrowband blue-light exposure has the potential to influence ocular growth and the diurnal ocular rhythms of eye length. Based on this information, we hypothesized that exposure to narrowband blue light would disrupt the normal rhythm of axial length change, which is tested as objective 1 in this study.

Thereafter, we conducted a second experiment to investigate whether narrowband blue light interacts with optical myopic defocus in humans. Current understanding of the combined effects of optical defocus and different narrowband lights is derived primarily from animal studies, ^4,7,11,17–26^ which show that the interaction between myopic defocus and blue light produces a greater reduction in axial length than blue light alone (See *supplementary Table 1*). In contrast to animal findings, human studies examining the short-term effects of induced myopic defocus alone have reported mixed results, with some studies reporting a small reduction in axial length and others reporting no significant effect. ^27^ With respect to blue light and myopic defocus, combining red-pass filters with an extended depth of focus contact lens was reported to increase the axial length, whereas combining blue-pass filters resulted in a shorter axial length.^13^ These findings are consistent with our previous observation, in which one-hour exposure to red light and green light led to an increase in axial length in both defocussed (hyperopic defocus) and fellow eye, but no such effect was observed following blue light exposure. However, uncertainty still exists regarding the interaction effect of myopic defocus and narrowband blue light on axial length. Considering the role of longitudinal chromatic aberrations, blue light effectively simulates a myopic (wavelength-dependent) defocus. Therefore, we hypothesized that the combination of optical myopic defocus and blue light exposure would produce a stronger short-term reduction in axial length compared with blue light exposure alone.

Based on this background, the present study comprised two experiments with the following objectives: (i) to investigate the effects of morning and evening narrowband blue light exposure on axial length, and (ii) to examine the interaction between narrowband blue light exposure and myopic defocus on axial length in humans. For the second objective, we chose the morning session where narrowband blue light exposure was found to exert the most influence to determine if further exposure with myopic defocus amplifies the effect. For both objectives, we had broadband white light exposure as a control.

## Methodology

The study protocol for both experiments was approved by the Institutional Review Board (LEC 05-19-256) of LV Prasad Eye Institute and adhered to the tenets of the Declaration of Helsinki. Optometry students and staff from the institute participated in the study. After a thorough explanation of the study’s nature and potential effects, each participant provided informed written consent. All participants demonstrated a best-corrected visual acuity of 0.0 logMAR or better while wearing their habitual refractive error correction (if any). The study recruited young adults with astigmatism of less than 1.25 diopter (D) and with no history of any systemic condition, such as diabetes, hypertension, any ocular pathology, ocular injuries/surgeries, or sleeping disorder/medications (verbally asked to participants), that could affect the experiment’s outcome. Myopia was defined as non-cycloplegic spherical equivalent ≤ -0.75 D, and non-myopes as ≥ -0.50 D measured with an open-field autorefractor (Shin-Nippon NVision-K 5001, Japan). For both objectives, the sample size was calculated using G*Power ^28^ based on the axial length change value used by Liu et al.(0.015 ± 0.02 mm).^13^ Using a paired t-test with a study power of 80%, the required sample size was determined to be 16 participants. A total of 18 participants were recruited for objective-1, and 27 participants were recruited for objective-2, both exceeding the required sample size.

### Experimental set-up

Our previous study detailed the experimental setup, which remained the same across both objectives.^29^ Narrowband blue light (peak at 460 nm, average irradiance 0.00448 W/nm/m^2^, and half maximum width = 35 nm, purity 98.3%) was used as the experimental light condition. Broadband white light with a prominent “blue” peak (average irradiance 0.00495 W/nm/m^2^, purity 17.4%) acted as a control, as shown in *Figure 1.* The light sources for both conditions consisted of 11 light-emitting diode smart bulbs (12-Watt, Wipro Enterprises Ltd., Shenzhen, People’s Republic of China, China) mounted in a room measuring 3.0 m length x 2.0 m width x 3.2 m height. The “Wipro Next Smart Home” and Google Home” smartphone application, integrated with Google Voice Assistant, was used to connect and synchronize all 11 bulbs. This setup allowed the examiner to switch the color of all the bulbs simultaneously. During the experiment, all bulbs were operated at full brightness (100%).

**Figure 1:**
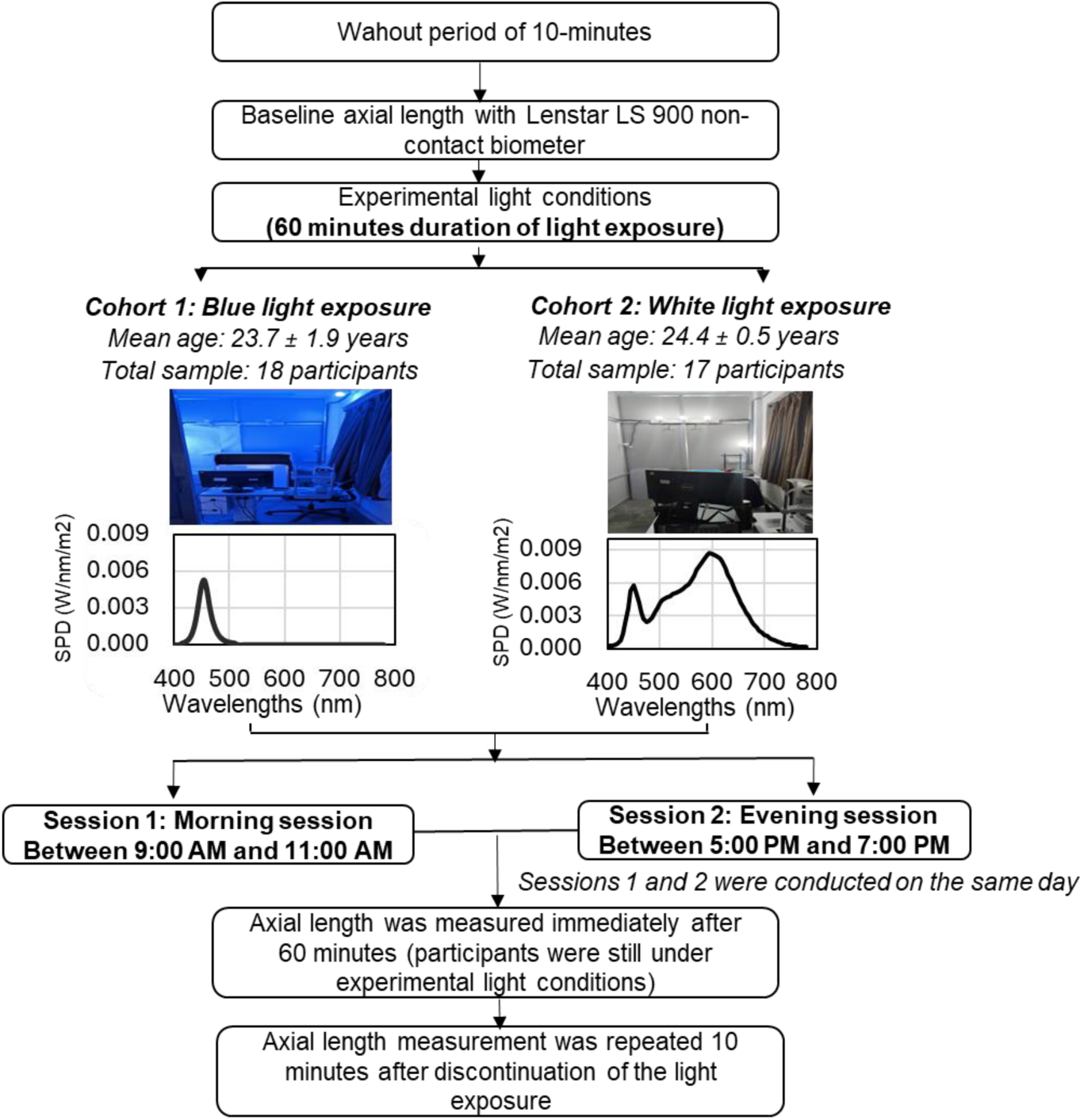
Flow diagram for the objective-1 methodology.

**Figure 2:**
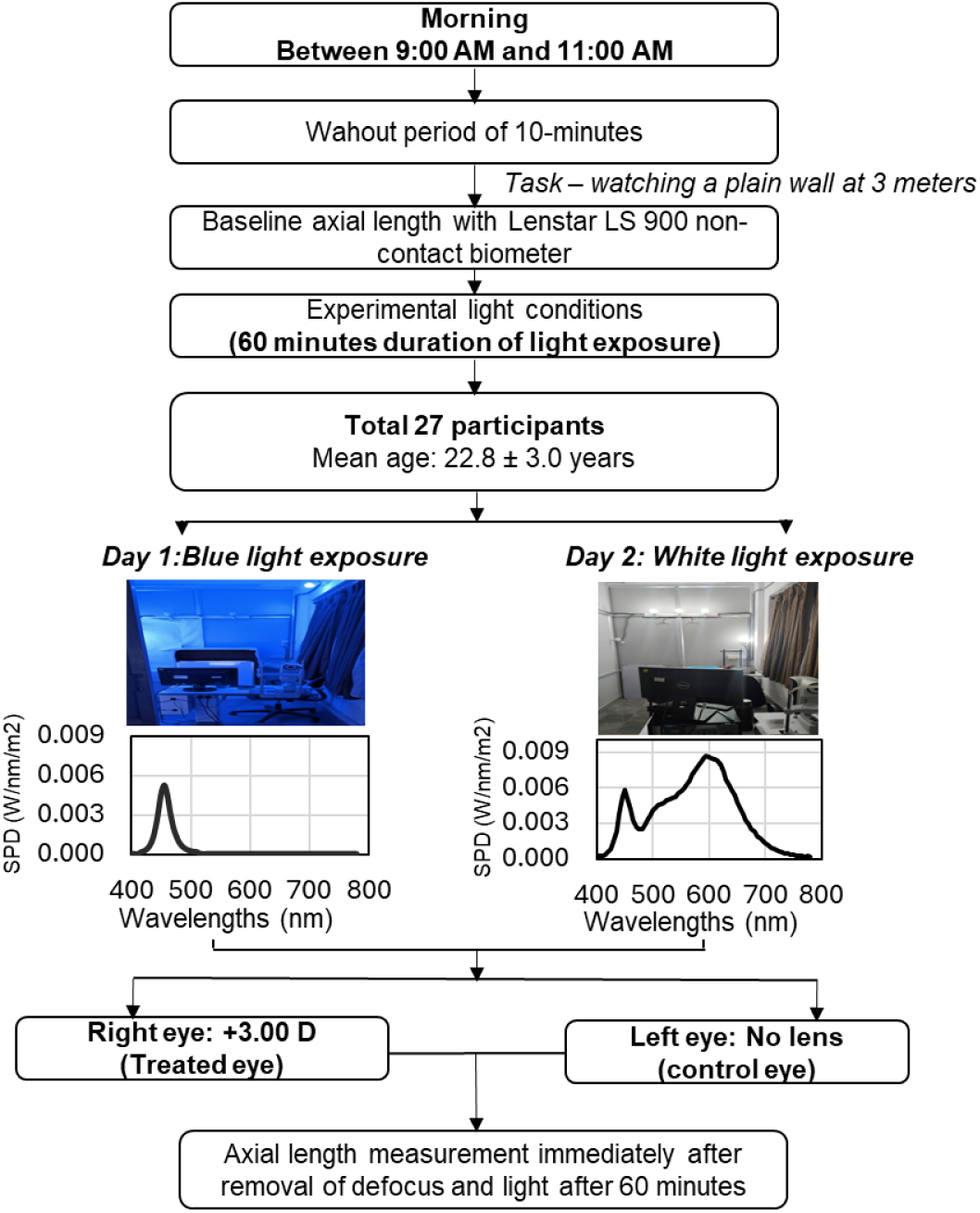
Flow diagram for the objective-2 methodology.

Participants watched a movie on a laptop (Apple MacBook Air 2017, screen size = 13.3 inches, and display resolution = 1440 x 900 pixels) positioned at a viewing distance of 3 meters from the eye during 60 minutes of exposure to narrowband blue light and broadband light exposure. To minimize spectral interference during blue light exposure, a blue-colored cellophane sheet was placed over the laptop screen. The peak transmission spectra of the blue cellophane sheet (450 nm) were similar to the narrowband blue light used in this experiment, ensuring a uniform narrowband light environment.^29^ No colored cellophane sheet was placed over the laptop during white light exposure. A hand-held portable spectrometer (Photonfy SP-01-BLU, Ledmotive, Spain) calibrated to the visual V-lambda was used to measure the spectral profile of the experimental illumination condition at the eye level. This pocket-sized spectrometer (with an Android application) allowed examiners to capture and record the light spectrum in real time (spectral range: 380-780 nm, maximum resolution, 10 nm).

Before the experiments began, each participant underwent a 10-minute washout period in a broadband white light condition, during which they were instructed to relax, avoid digital devices, and look at a distance with no specific fixation target. This resting period aimed to minimize the effects of prior near-work activities on axial length. After the washout period, baseline axial length measurements were conducted under broadband white light. Once the baseline measurement was completed, the narrowband blue light was turned on to begin the experimental exposure. Lenstar LS-900 non-contact biometer (Haag Streit AG, Koeniz, Switzerland) was used to measure the axial length, with an average of five biometric readings considered for the analysis. To prevent the biometer’s white flash of light from influencing the outcomes, the white-to-white analysis for the instrument was turned off during the experiment. The biometer was positioned close to the participants to minimize the movement.

### Objective-1: Effect of morning and evening narrowband blue light exposure on axial length

A total of 18 participants with a mean age of 23.7 ± 1.9 years (10 emmetropes and 8 myopes) were recruited for both morning and evening narrowband blue light exposure. For the broadband white light exposure (control), we were unable to recruit the same cohort due to logistical constraints and hence recruited age- and axial length-matched 17 participants with a mean age of 24.4 ± 0.5 years (10 emmetropes, 7 myopes; mean axial length: 23.54 ± 0.36 mm; range: 21.55–27.17 mm). An independent samples t-test showed no significant difference in age (p = 0.39) and axial length (p = 0.23) between the narrowband blue light and control cohorts.

The protocol for either narrowband blue light exposure or broadband white light exposure included two experimental sessions conducted on the same day: morning (9:00 AM to 11:00 AM) and evening (5:00 PM to 7:00 PM), *Figure 1*. Each session included 60 minutes of light exposure. Briefly, following the 10-minute washout period, baseline axial length measurements were obtained under broadband white light, following which either the blue light or white light was turned on for 60 minutes. The measurements for both eyes were repeated after 60 minutes of light exposure (participants were still under experimental light conditions) and 10 minutes of discontinuing narrowband blue light or broadband white light under normal room light conditions to assess the change in axial length after light exposure (i.e., at 70 minutes). The rationale for performing the measurements 10 minutes after discontinuation was to evaluate the effect of the experimental light exposure on the daily rhythm of axial length, rather than capturing only the immediate light-induced response. Since the measurement was conducted first on the right eye, only data from the right eye were included in the analysis for this study. The same procedure was followed during morning and evening blue light sessions. During both light conditions, participant with myopic correction wore their best-corrected single-vision spectacle correction. The mean axial length for the right eye was 24.16 ± 0.37 mm (range, 21.77 mm to 26.84 mm). The mean ± SD for the right eye was -1.42 ± 0.62 D.

### Objective-2: Interaction between morning narrowband blue-light exposure and myopic defocus on axial length

A total of 27 participants with a mean age of 22.8 ± 3.0 years (20 emmetropes, 7 myopes) visited the lab on two separate experimental days in the morning (9:00 am to 11 am). On both days, the experiment was conducted between 8:00 am-11:00 am, where the participants were exposed to a constant 60-minute duration of either narrowband blue light or broadband white light. The session of narrowband blue light and broadband white exposure was randomized across participants. During both light conditions, participant with myopic correction wore their best-corrected single-vision spectacle correction, with an extra +3.00 DS trial lens placed in front of the right eye using a trial frame clip-on (large aperture) to impose full-field myopic defocus for 60 minutes. Emmetropic participants wore a trial frame with a +3.00D trial lens in front of the right eye and a plano trial lens in front of the left eye. The trial lens, which was used to impose myopic defocus, did not prevent the transmission of narrowband blue light. The right eye always served as a treated eye (i.e., observing the combined effect of blue light and lens-induced myopic defocus on ocular biometry), and the left eye is the control eye (i.e., to evaluate the effect of blue light or white light alone). A similar paradigm of monocular defocus has been applied in previous primate and human experiments reporting the interaction of optical defocus and wavelength.^27,30^

The biometry measurements were conducted before and after 60 minutes of narrowband blue and white light exposure. The measurements were always performed first on the right eye (treated eye experiencing both optical defocus and light exposure) immediately after removing the spectacles or trial frame, with both eyes measured within one minute of defocus removal. The data for both the right eye (treated eye) and left eye (control eye) were considered for the analysis. The mean (standard deviation, SD) axial length for the right eye and left eye was 23.48 ± 0.93 mm (range, 22.20 mm to 25.61 mm) and 23.46 ± 0.96 mm (range, 22.13 mm to 25.56 mm), respectively. The mean ± SD SER (spherical equivalent refraction) was -0.38 ± 0.55 D and -0.30 ± 0.50 D for the right and left eye, respectively.

To assess the impact of combining defocus with blue light on axial length, following comparisons were made for changes in the axial length: eye with defocus and blue light (right eye data) versus eye with defocus and white light; eye with defocus and blue light (right eye data) versus eye with blue light exposure only (left eye data), eye with defocus and white light (right eye data) versus eye with white light exposure only (left eye data from same day/session). There was no overlap of participants between objective-1 and objective-2.

## Statistical analysis

All statistical tests were conducted using IBM SPSS Statistics version 21.0.0 (SPSS, Inc., Chicago, IL, USA). The normality test was performed using the Shapiro-Wilk test, which revealed a normal distribution for changes in axial length (p>0.05). With regards to objective-1, a paired t-test was used to evaluate the change in axial length from baseline for both morning and evening sessions. To highlight the diurnal variation, the axial length values at each time point were compared to the normalized axial length, i.e., the average of the morning and evening axial length values separately for values obtained before and after light exposure. In objective-2, a 3-factor repeated measure ANOVA (defocus, time, and light condition) was performed to identify the main effect of each one of these factors on the change in axial length, and a paired t-test was employed for interocular comparisons. An independent t-test was conducted to detect the statistical significance of changes in axial length between emmetropes and myopes. The change in axial length values was represented as mean ± standard error of mean (SEM), and a p-value of <0.05 was considered statistically significant.

## Results

### Objective-1: Effect of morning and evening narrowband blue light exposure on axial length

#### Change in axial length from baseline

*Figure 3* shows the change in axial length from baseline after narrowband blue and broadband white light exposure. After 60 minutes, when participants were still under the experimental light condition, a similar magnitude (p=0.67) of reduction in axial length was noted for both morning (-6.11 ± 4.21 µm) and evening (-7.77 ± 4.08 µm) blue light exposure (*Figure 3a*). For the measurements obtained ten minutes after completion of light exposure (i.e., 70 minutes), there was a significant reduction in axial length for morning narrowband blue light but not with evening blue light exposure (morning: −10.00 ± 3.96 µm vs evening: −0.67 ± 3.30 µm; p = 0.02), whereas no such effect was observed with broadband white light exposure (0.0 ± 3.54 µm vs. -2.50 ± 4.23 µm, p = 0.70). Individual changes in axial length for all participants after completion of narrowband blue light and broadband white light exposure during morning and evening are shown in *Supplementary Figure S1*. Emmetropes and myopes showed similar effect in changes in axial length after both morning and evening light exposure (in both 60 minutes and 70 minutes measurements), as shown in *Table 1*.

**Figure 3:**
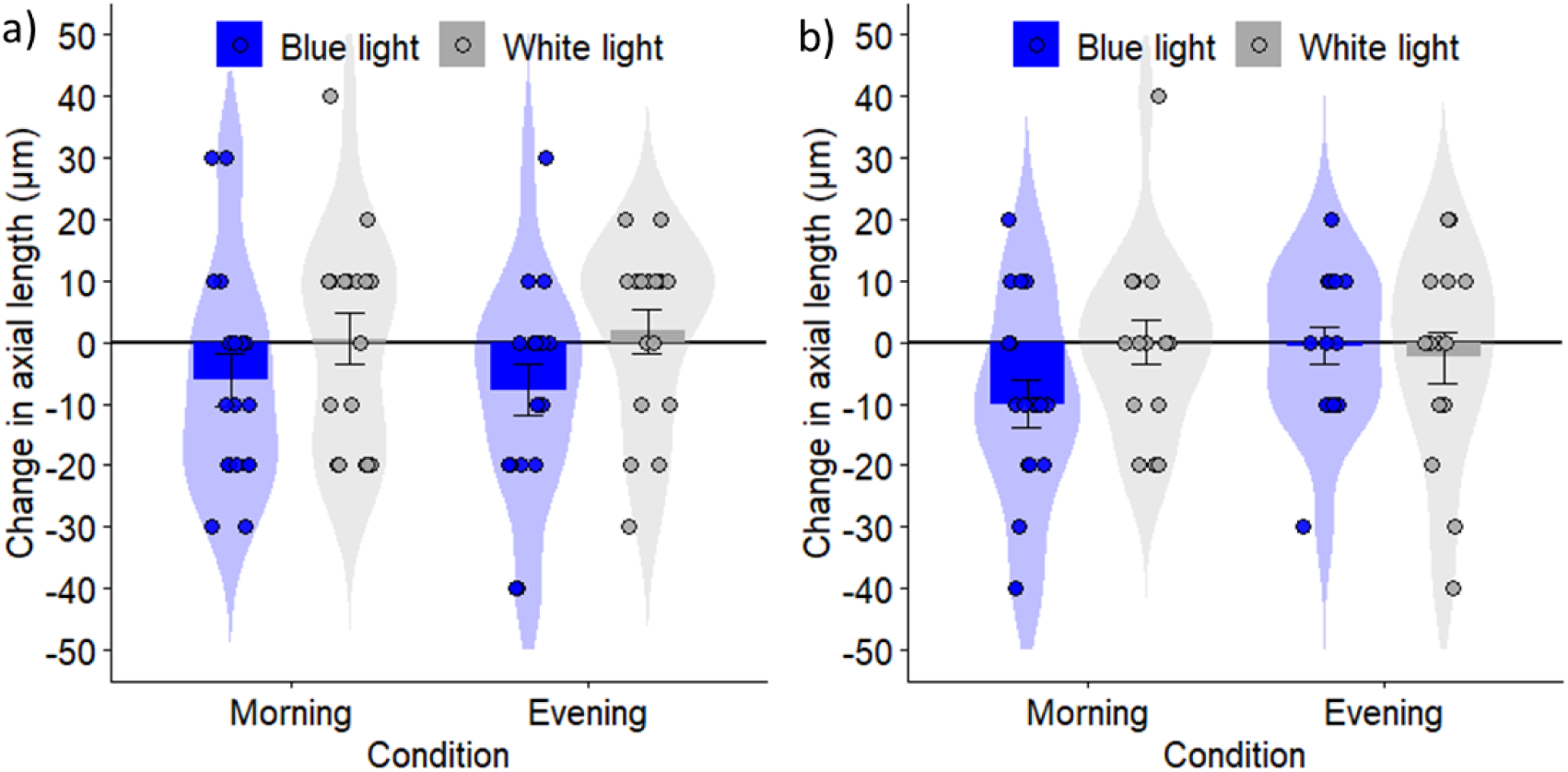
Change in axial length in participants exposed to narrowband blue light and broadband white light after a) 60 minutes, and b) 10 minutes of discontinuation of light exposure.

**Table 1:** Mean ± standard error of mean change in axial length in overall, emmetropes, and myopia at 60 minutes and 10 minutes after discontinuation of light exposure (i.e., 70 minutes).

| Wavelength | Time of day | 60 minutes (Immediate response) |  |  | After 10 minutes of discontinuation |  |  |
| --- | --- | --- | --- | --- | --- | --- | --- |
|  |  | Overall | Emmetropes | Myopes | Overall | Emmetropes | Myopes |
| **Blue light** | Morning | -6.11 ± 4.21 | -7.00 ± 5.97 | -5.00 ± 6.26 | -10.0 ± 3.96 | -10.00 ± 4.47 | -10.0 ± 7.32 |
| | Evening | **-7.77** ± 4.08 | -7.00 ± 6.15 | -8.75 ± 5.48 | -0.67 ± 3.30 | +3.75 ± 3.24 | -5.71 ± 5.71 |
| **White light** | Morning | +0.58 ± 4.24 | +2.72 ± 5.23 | -3.33 ± 7.60 | 0.0 ± 3.54 | 0.00 ± 4.86 | 0.0 ± 5.16 |
| | Evening | +1.76 ± 3.55 | -2.72 ± 4.87 | +10.0 ± 2.58 | -2.50 ± 4.23 | -7.00 ± 5.78 | +5.00 ± 5.00 |

#### Effect on diurnal variation

*Figure 4* illustrates the differences in axial length across time and with light exposure. The normalised baseline axial length measurements revealed that the axial length was shorter in the evening than morning for both the groups before light exposure (narrowband blue light group: –7.06 ± 2.23 µm versus +1.76 ± 1.76 µm, p = 0.04, broadband light exposure group: –7.06 ± 2.23 µm versus -0.58 ± 2.00 µm, p = 0.13). After light exposure, the change in axial length (compared to the normalized values) was similar to that of before light exposure in the broadband white light group (*Figure 4b:* morning: +2.35 ± 1.82 µm vs. evening: –6.25 ± 2.21 µm, p = 0.04). However, such a trend was not observed after the blue light exposure group, with no statistical differences in the morning and evening axial length (*Figure 4a:* morning: -4.12 ± 1.72 µm vs. evening: -2.00 ± 2.00 µm, p = 0.48).

**Figure 4:**
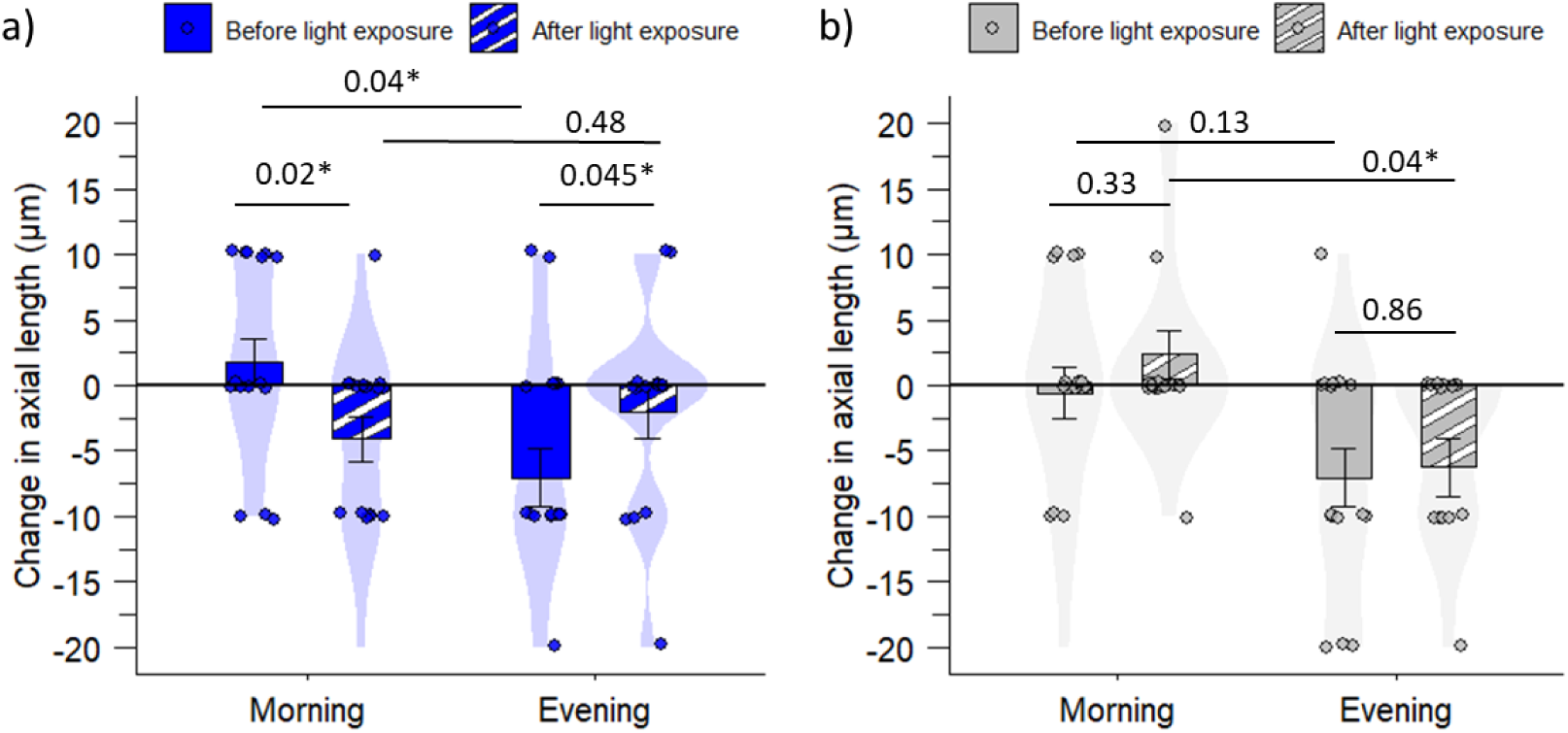
Diurnal changes in axial length are shown for before and after exposure to a) narrowband blue light and b) broadband white light. Error bars represent the standard error of mean (SEM).

Further, morning blue light exposure led to a significant shortening of axial length (before exposure: +1.76 ± 1.76 µm vs. after exposure: −4.12 ± 1.72 µm; p = 0.02). In contrast, evening blue light exposure resulted in a smaller change compared with before exposure (before exposure: −7.06 ± 2.23 µm vs. after exposure: −2.00 ± 2.00 µm; p = 0.045). Such alterations were not observed with broadband white light exposure (morning: p = 0.33; evening: p = 0.86).

### Objective-2: Interaction between narrowband blue-light exposure and myopic defocus on axial length

The combined effect of defocus and light exposure was considered only for the morning session. The change in axial length with myopic defocus combined with blue light exposure (treated eye) and only blue light (control eye) were -2.22 ± 3.26 µm and −3.70 ± 3.16 µm, respectively; corresponding values with myopic defocus under broadband white light (treated eye), and only broadband light (control eye) were -0.37 ± 3.90 µm, and -4.07 ± 2.34 µm, respectively (*Figure 5)*. The effect of defocus (treated eye vs. control eye), light exposure (change from baseline), and type of light condition (narrowband blue and broadband white light) in a 3-factor RM-ANOVA revealed no significant effect of myopic defocus (F_(1,26)_ = 0.28, *p* = 0.59), time (F_(1,26)_ = 1.54, *p* = 0.22), and the type of light condition (F_(1,26)_ = 0.15, *p* = 0.90). The differences in axial length between the two eyes (interocular), for both narrowband blue light and broadband white light exposure, and between light conditions (in both with and without defocus conditions) were not statistically significant. Both the emmetropes and myopes showed a similar trend in axial length after narrowband blue light (emmetropes: -1.00± 4.16 µm; myopes: -5.71 ± 4.28 µm, p=0.54) and broadband white light exposure (emmetropes: +2.00 ± 4.67 µm; myopes: -7.14 ± 6.80 µm, p=0.31); outcomes were similar in the control eyes.

**Figure 5:**
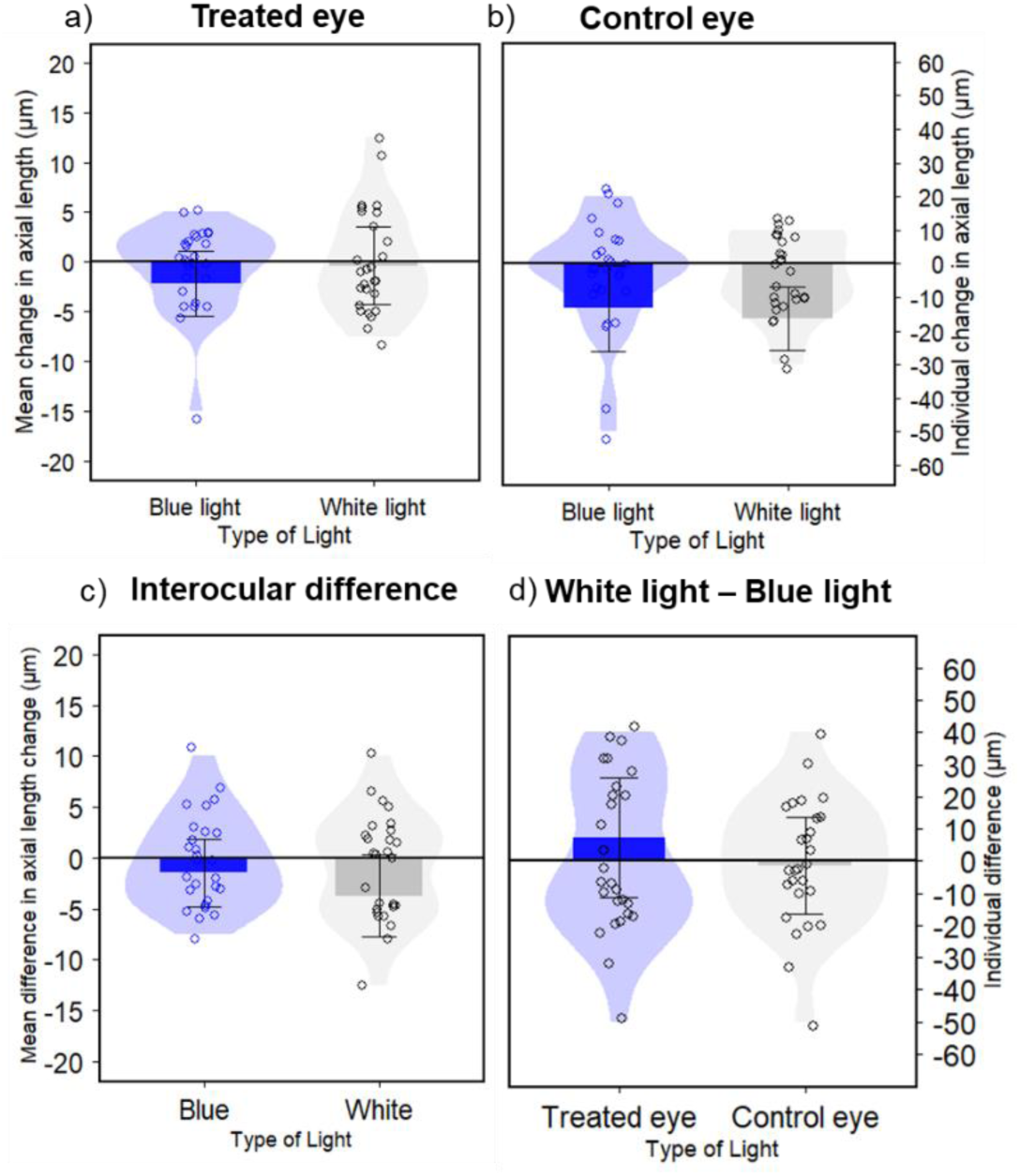
a) Mean change in axial length after narrowband blue light and white light exposure in the a) treated eye (right eye), b) control eye (left eye), c) Mean difference in change in axial length between the treated eye and the control eye (right eye - left eye) and d) mean difference in change in axial length between broadband white light and narrowband blue light exposure. The primary y-axis shows the mean change (bar plot), and the secondary y-axis (violin plot) represents the individual change in the axial length.

## Discussion

The current study addressed two questions regarding the impact of narrowband blue light on axial length: (i) the effect of morning and evening narrowband blue-light exposure, and (ii) the interaction between blue light and lens-induced myopic defocus. The first objective demonstrated that morning exposure to narrowband blue light attenuated the diurnal rhythm, resulting in a greater reduction in axial length during the morning than in the evening; no comparable effect was observed with broadband white light exposure. When the protocol was repeated for the second time, only in the morning to evaluate the combination of light and defocus, the impact of blue light alone was not different to that of broadband white light, and lens-induced myopic defocus did not result in a significant change in axial length, regardless of the light condition.

### Objective-1: Effect of morning and evening narrowband blue light exposure on axial length

Our hypothesis that morning exposure to narrowband blue light can alter the diurnal variation of axial length in humans is primarily supported by evidence from other fields of light research. Its role in modulating daily cycles of metabolic function,^31^ brain activity, and cognitive performance ^32^ and melatonin and cortisol levels ^33^ has been reported previously. To our knowledge, this study provides new human evidence on the time-dependent effect of narrowband blue-light exposure on diurnal variation in axial length. Our findings highlight that even a low intensity of narrowband blue light for a shorter duration could alter the daily rhythm and diminish the morning-evening difference of axial length (normal rhythm). These outcomes closely relate to the findings reported by Liu and colleagues. ^13^ investigating the time-dependent effects of combining extended depth of focus (+2.50 D) with blue-pass filters (cut-off wavelength beyond 530 nm) and red-pass filters (cut-off wavelength shorter than 580 nm) at two different time-points: morning (09:00 and 11:00 am) and afternoon (15:00 and 17:00 pm). Their findings indicated that instead of elongated axial length during morning, blue-pass filters attenuated the normal rhythm of axial length (morning: 0.002 ± 0.003 mm; evening: -0.004 ± 0.003 mm), while the red-pass filters showed a small increase in axial length both in the morning (+0.013 ± 0.002 mm) and in the afternoon (+0.006 ± 0.002 mm). Extending these findings to animal studies, Nickla et al.^10^ reported that 4 hours of narrowband blue light exposure at low illuminances (<1000 lux) around the transition period of the day stimulated ocular growth and disrupted axial length rhythms in chicks, suggesting that morning or evening blue light exposure may influence eye growth and myopia development, unless exposure occurs at higher illuminances (≈1000 lux or above), as typically encountered outdoors. But the influence of narrowband light on ocular growth is considered to arise from a complex interaction not only with light intensity but also likely with duration of exposure, wavelength specificity, time of day, circadian rhythm, hormonal regulation, bandwidth, temporal sensitivity, and prior light exposure. ^34–37^ Given the short-term nature of these experiments, it is challenging to fully control the influence of these factors on the outcomes, particularly in experiments involving human participants. As the evidence on the effects of narrowband blue light on ocular biometric parameters in humans remains limited and evolving, further investigations examining the time-of-day dependent effects of narrowband blue light on continuous 12-hour and 24-hour cycles on axial length are warranted to reveal whether its phase-advances or delays the rhythm. Additionally, these findings are relevant to the ongoing discussion about whether extensive use of digital devices may contribute to an increased risk of myopia in children. ^38,39^ Smartphones and other handheld screens now constitute a substantial component of children’s daily activities and emit significant amounts of blue-enriched white light. Beyond the closer viewing distances and increased accommodative demand they impose, one plausible pathway through which smartphone use may influence ocular growth is by altering the normal diurnal rhythm of axial length in children, which needs to be tested in the future.

### Objective-2: Interaction between narrowband blue light exposure and myopic defocus on axial length

Contrary to our expectation, the 60 minutes of myopic defocus of 3.00 D did not influence the axial length, neither in the broadband white light nor in the narrowband blue light condition. Other short-term experiments in young adults and children have indicated a small mean reduction in axial length after 60-120 minutes of exposure to monocular myopic defocus (Read et al: -13 ± 14 µm with +3.00 D for 60 min; Delshad et al.: −8_±_10 μm with +3.00 D for 60 min; Wang et al: -5.88 ± 6.19 µm in 60 min and -14.12 ± 7.53 µm with +3.00 D in 120 min; Moderiano et al: -0.021 ± 0.009 mm with +3.00 D in 120 min; Swiatczak and Schaffel: -8.8 ± 9.2 µm with +2.5 D for 60 min).^40–44^ Note that across these studies, a significant heterogeneity could be observed, suggesting that not all individuals responded similarly to defocus. This, in fact, is consistent with our observation where a certain percentage of individuals showed an expected reduction in axial length, and others showed an opposite trend, or no change in axial length. While the exact reason for such heterogeneity is difficult to deduce based on the current experimental paradigm, there could be a few possible reasons that could be considered, such as the i) presence of active accommodation during the experiment, which can dampen the magnitude of blur experience at the retinal level, ^45^ ii) ciliary muscle contraction resulting in thinner choroid or less axial length change ii) retinal illuminance of blue light due to pupillary miosis, and iii) and loss of spatial information required for regulating ocular growth,^46–48^ and iv) refractive status of the individual. Swiatczak and Schaffel attributed the variation in response to myopic defocus to the refractive status of the individuals, where emmetropic participants showed a significant reduction in axial length, while individuals with myopia exhibited no effect of myopic defocus. ^44^ However, we noted that 11 out of 20 emmetropes showed either an increase or no change in axial length in response to myopic defocus, and the remaining 8 showed a reduction in axial length in response to myopic defocus under broadband light conditions. While we acknowledge the report by Swiatczak and Schaffel on how the response to stimuli by emmetropic and myopic eyes could be different, it is beyond the scope of this paper to explain the underlying reason for no differences between the refractive groups.

Additionally, the magnitude of axial length shortening following exposure to narrowband blue light was greater when blue light was presented alone (objective-1) compared to when it was combined with monocular myopic defocus (objective-2). This attenuation may reflect non-linear integration of retinal growth-inhibitory cues at the retinal level, where competing signals do not show a linear summation effect, but instead exhibit saturation, dominance, or re-weighting of visual inputs ^49^, which may result in a sub-additive or even reversal axial length response. Supporting this interpretation, Ingrassia et al.^50^ demonstrated in human participants that combining two visual stimuli that individually induced axial shortening did not produce an additive effect; rather, the combination of an inverted contrast-polarity target with a red-in-focus filter resulted in axial length elongation compared to inverted contrast alone, which produced axial shortening. On a similar note, in chickens, Sarfare et al.^51^ demonstrated that the effect of bright light exposure on ocular growth differs when combined with optical defocus, compared to normally growing eyes. In addition to these factors, methodological considerations such as reduced retinal blue light irradiance due to the presence of a trial frame or lens/spectacles lens may have further contributed to the attenuated effect of narrowband blue light on axial length.

### Individual differences in response to short-term light exposure

Despite differences in the experimental paradigms between objective-1 and objective-2, narrowband blue light exposure also showed significant inter-individual variability in response. Some participants showed a significant change in axial length (increase or decrease), while others showed no measurable change in axial length. These inter-individual differences were observed irrespective of refractive error. Beyond ocular biometric parameters, Spitschan and Santhi have reported that inter-individual differences in light responses are also evident in other physiological parameters in healthy human participants.^52^ For example, the most sensitive individuals experienced 50% melatonin suppression at an illumination of 10 lux (approximately similar to dim reading light illumination), whereas the least sensitive individuals required 40 times brighter light for the same level of suppression.^52,53^ Similar variability has been demonstrated for pupil response to light.^54^ Several potential factors have been proposed to account for these individual differences, including i) retinal illuminance may vary between individuals, ii) health status and use of medication, iii) genetic variability affecting the sensitivity of melanopsin signalling characteristics, iv) individual differences in spectral sensitivity, and v) photic history.^52^ While we have recruited the healthy young adults for both experiments, it is beyond the scope of this study to explain the reasons for such variability.

### Limitations

The findings of this study should be interpreted in light of several limitations. First, for the first objective, the control participants were not the same individuals as those in the blue-light group; however, the groups were matched for age and baseline axial length to minimise potential confounding effects. Secondly, the 12-hour or 24-hour diurnal variation was not mapped for the participants, which might reveal a better understanding of the effect of narrowband blue light exposure on the diurnal variation of axial length. The second objective was limited to a single strength of full-field myopic defocus (+3.00 D) and included only narrowband blue light, with broadband light serving as the control; exposure to long-wavelength light was not examined. Consequently, different outcomes may be expected with a greater magnitude of defocus, the inclusion of peripheral defocus, higher light intensities, longer duration of exposure, or other wavelengths. Individual differences in light sensitivity (i.e., the same light may affect individuals differently) and prior light exposure were not taken into account in either experiment. Nevertheless, the duration, intensity, and spectral composition of the narrowband blue light were kept constant across both objective-1 and objective-2. Lastly, the use of a monocular defocus paradigm, similar to that employed in previous experiments, may partly explain the absence of an effect of myopic defocus on axial length, as participants may have preferentially used visual input from the unblurred eye under binocular viewing conditions.

## Conclusion

In conclusion, the short-term narrowband blue light exposure in humans exerted a time-of-day-dependent effect on axial length, with a small but significant decrease in axial length from baseline in the morning compared to evening exposure. The blue light exposure diminished the usual diurnal rhythm in axial length, whose implications for regulating ocular growth in long-term to be tested further. The combination of blue light with lens-induced myopic defocus did not provide additional short-term modulation of axial length, which needs to be further examined with a higher magnitude of defocus, or the existing myopia control defocus lenses and brighter blue light intensities, to improve our understanding of the potential application of combining light therapy with defocus lenses in myopia control.

## Data Availability

The data will be provided based on the appropriate request.

## Acknowledgement

We also want to acknowledge Mr. Rakesh Maldoddi for helping us to conduct data collection for broadband white light exposure for objective-1.

## Commercial Relationships Disclosure

op*&?”-;[6.

