## Supplemental Figure 1 for "Effects of morning and evening narrowband blue light and myopic defocus on axial length in humans"

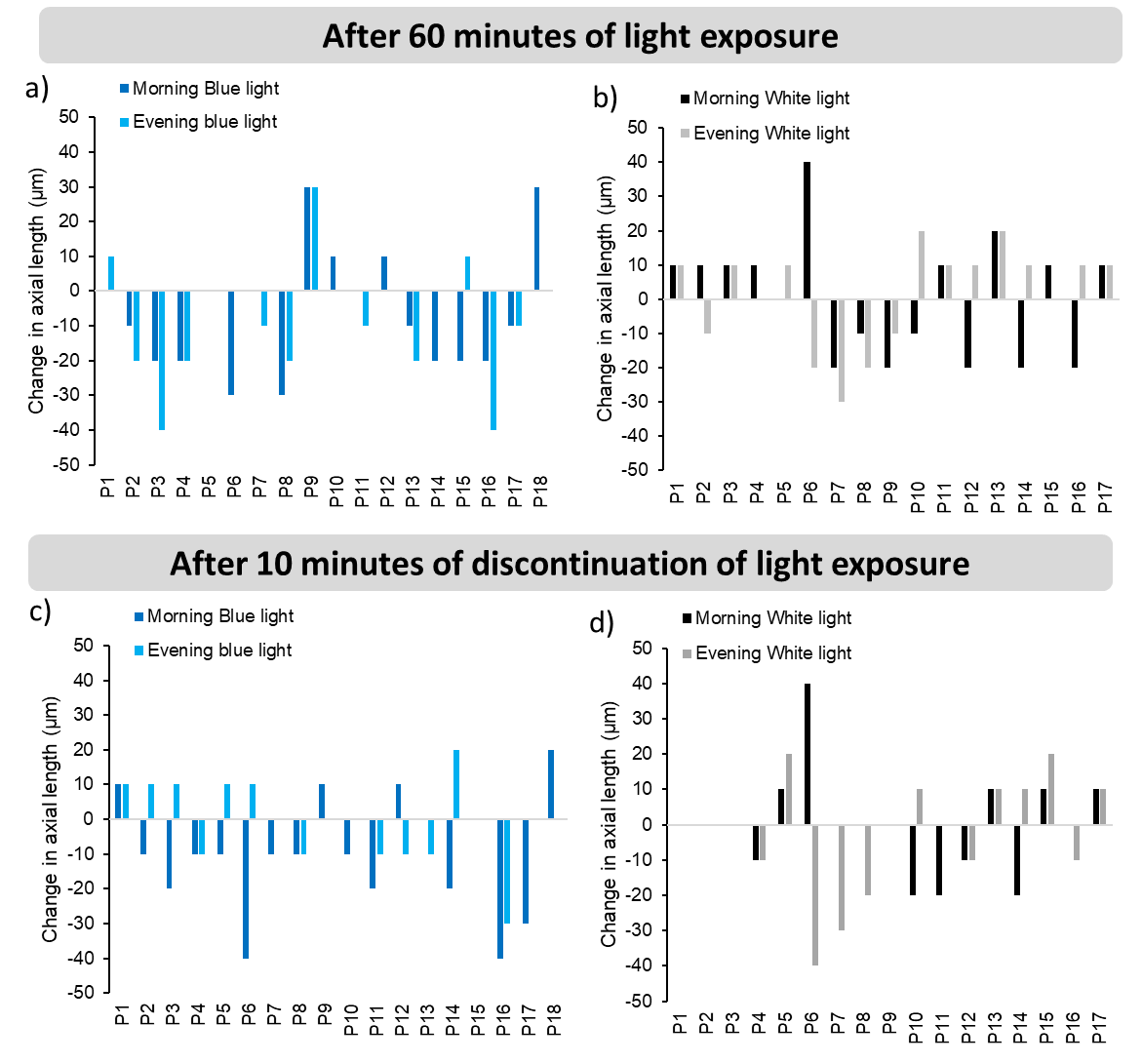

***Supplementary Figure S1:*** *Distribution of change in axial length under morning and evening a) narrowband blue light (P1-P18), and b) broadband white light exposure (P1-P17) after 60 minutes, and 10 minutes of discontinuation of light exposure. *if the bar is not visible, then it indicates 0 change in axial length.*
