## Supplemental Table 1 for "Effects of morning and evening narrowband blue light and myopic defocus on axial length in humans"

| *Supplementary Table 1 summarizes both animal and human studies reporting the time-dependent effect of different wavelengths of light, and the interaction effect of different wavelengths of light and optical defocus on ocular growth* | | | | | |
| --- | --- | --- | --- | --- | --- |
| *Author (year)* | ***Species*** | ***Visual manipulation*** | ***Wavelengths (nm)*** | ***Intensity/Duration*** | ***Outcomes*** |
| Objective-1: Effect of morning and evening narrowband blue light exposure on axial length | | | | | |
| Nickla et al. (2022)^1^ | Chicks | - | Narrowband blue light: 460 ±10 nm | Evening exposures: 0.15 lux, 200 lux, 600 lux, 1000 lux for 4 h/day (9 days); 600 lux also tested for 2 h/day; Morning exposure: 200 lux, 600 lux, 1000 lux for 4 h/day (9 days) | Evening blue light (≤ 600 lux) stimulated ocular compared with white light; 1000 lux showed no effect. Morning 200 lux exposure increased ocular growth. Time-of-day dependent effect was observed |
| Liu et al. (2025) | Young adults | **-** | blue-pass, red-pass | Room illumination 600-620 lux; 1 hour | Blue-pass filters mitigated the normal diurnal rhythm in axial length |
| Current Study | Young adults | **-** | Blue light (460 nm) | Blue (0.000174 W/nm/m2); 1 hour | Morning exposure to narrowband blue light disrupted the diurnal rhythm of axial length. Broadband white light exposure did not have any effect on the diurnal rhythm of axial length |
| Objective-2: Interaction between narrowband blue light exposure and myopic defocus on axial length | | | | | |
| Rucker and Wallman (2008)^2^ | Chick | ±6D, ±8D lenses | Blue light (460 nm) and red light (620 nm) | 0.67 lux; day-weeks | Under hyperopic defocus, blue light showed an increase in axial length compared to red. With myopic defocus, a greater reduction in axial length was noted with blue light compared to red light. |
| Jiang et al. (2014)^3^ | Guinea pigs | +4 D and -4 D | Red light (600 ± 5), Blue light (470 ± 5), and Broadband light (6500 K) | 50 lux for blue, 300 lux for red, and 350 lux for white; 4 weeks | Blue light inhibited the effect of hyperopic defocus, and red light had no effect when combined with myopic defocus |
| Hung et al. (2018)^4^ | Rhesus monkey (infant) | +3 D/-3 D, diffuser | Red light (630 nm) | 274 + 64 lux; 12 hrs/day | Red light promoted hyperopia in the presence of hyperopic defocus and form-deprivation. Same trend in myopic defocus |
| Strickland et al. (2020)^5^ | Guinea pigs | -10 D | Violet (400 ± 20), Green (525 ± 40), and broadband (420-680) | 50 cd/m2; 4 weeks (12:12 cycle) | Violet light suppressed the effect of hyperopic defocus, whereas green and white light resulted in a myopic shift in the presence of hyperopic defocus |
| Jiang et al. (2021)^6^ | Mice | -30 D | Violet light (360 to 400), Blue light (440 to 480), Green light (500 to 540), Red light (610 to 650) | ~400 µw/cm^2^; 3 hrs/day | Red and green lights did not show an inhibitory effect. Violet light had a stronger inhibitory effect on myopic defocus, followed by blue light |
| Yu et al. (2021)^7^ | Guinea pigs | -5.00 D | Blue light (440 nm) and white light | 500 lux for blue and 580 lux for white; 4 weeks | Blue light inhibited the effect of hyperopic defocus |
| Riddell et al. (2021)^8^ | Chick | -10 D and +10 D | Long-wavelength filter B410 and B460 | 201 lux; 1 week | B410 (but not B460) long-wavelength–filtered light had significantly inhibited negative lens-induced axial growth |
| Nickla et al. (2023)^9^ | Chicks | -10 D | Blue light (460 ± 10) | 200 lux (dim) and 600 lux (bright); morning vs. evening (7 days) | Blue light enhanced axial compensation during evening time, and bright blue light in the morning showed an inhibitory effect compared to dim blue light |
| Jeong et al. (2023)^10^ | Mice | -30 D | Violet light (360 to 400) | Transmittance: 40%, 70%, 100% | The effect of hyperopic defocus was suppressed by violet light |
| Chun et al. (2023)^11^ | Chick | Dual-power optical lens (- 10 D/ + 10 D, 50∶50) | Red (634 ± 15), blue (451 ± 15), and white light | 250 lux; 12 hr/day for 2 weeks | Blue light plus dual-power lens had the shortest VCD and axial length, and red light did not show any significant effect compared to white light |
| She et al. (2024)^12^ | Tree shrew | -5 D, diffuser | Narrowband red (624 ± 10 or 634 ± 10) | 527-749 lux; 12-14 days (14 hr/day) | Red light reduced axial elongation and produced hyperopia, and slowed lens-diffuser-induced myopia |
| Ding et al. (2025)^13^ | Guinea pigs | -6.00 D | far-red/near-infrared light (710 ± 20) | 1.67 mW/cm^2^; 3 weeks | FR/NIR can effectively suppress myopia progression in LIM guinea pigs |
| Xian et al. (2025)^14^ | Guinea pigs | diffuser | Low-level red-light therapy (650) | 2.23 mW/cm^2^; 3 min per session with 4 hr interval for 3 weeks | Red light therapy retards FDM |
| Yu and Wildsoet^15^ | Chicks | +10/-10 D Fresnel lens | White light and blue light (460 nm) | 60 chick lux | Fresnel lens groups showed hyperopic shifts in refractive error, more so in blue light conditions |
| Human experiment | | | | | |
| Thakur et al. (2021)^16^ | Young adults | Hyperopic defocus (-3.00 D) | Red light (623), green light (521), blue light (460), and broadband white light | Red (0.00013 W/nm/m^2^), Blue (0.000174 W/nm/m2), Green (0.00021 W/nm/m2); 1 hour | An increase in axial length was noted for red and green light in the presence of hyperopic defocus, but not with blue light. No significant change in the choroid |
| Liu et al. (2025)^17^ | Young adults | Extended-depth-of-focus (+2.25 D) | blue-pass, red-pass, and neutral density filters | Room illumination 600-620 lux; 1 hour | A combination of blue-pass filters and myopic defocus showed an overall significant protective effect against axial elongation. No significant change in the choroid in any condition. |
| Hussain et al. (2026)^18^ | Young adults | Hyperopic defocus (-3.00D) | Cyan light (507 nm), Broadband white light | Cyan light (irradiance 3.06 W/m^2^), and broadband light (3.05 W/m^2^) | Cyan light reduced the axial length in the presence of hyperopic defocus |
| Current study | Young adults | Myopic defocus (+3.00 D) | Blue light (460 nm) and broadband white light | Blue (0.000174 W/nm/m2); 1 hour | No significant effect of myopic defocus on axial length in both broadband white and narrowband blue light conditions |
